# Multimodal Seizure Detection with Long-term Ambulatory ECG and Accelerometry Data

**DOI:** 10.64898/2025.12.16.25342428

**Authors:** Jieying Li, Philippa J. Karoly, David B. Grayden, Mark J. Cook, Ewan S. Nurse

## Abstract

**Objective:** Detecting epileptic seizures in real-world environments remains challenging, as electroencephalography (EEG) is often impractical in chronic ambulatory monitoring. Heart rate and accelerometry, measurable from wearable devices, provide a less obtrusive alternative. Although some studies explored multimodal wearable-based seizure detection, few have been validated on long-term ambulatory datasets reflecting real-world variability. This study investigated the added value of accelerometry for electrocardiography (ECG)-based seizure detection using a long-term, ambulatory video-EEG dataset. Using an established pseudo-prospective framework, we assessed performance across heart rate variability (HRV), accelerometry (ACC), and combined feature sets, and examined how it varied across patient groups with different seizure types.

**Approach:** Recordings from 78 patients (587 seizures, 385 days) undergoing ambulatory monitoring were analyzed. Time and frequency domain features were extracted from ECG and triaxial ACC. A logistic regression classifier trained on data from 47 patients was evaluated on a hold-out set of 31 patients. Seizure types were derived at the patient level through automated keyword extraction from clinical reports, enabling performance comparison across seizure type groups.

**Main results:** A combined HRV+ACC model achieved the best performance in leave-one-patient-out validation (Improvement over chance AUC, i.e., ΔAUC=0.12), while the HRV-only model generalized best to the hold-out set (ΔAUC=0.046). Across feature sets, 74% of patients demonstrated better-than-chance performance. Patients diagnosed with focal epilepsy, and those with substantial seizure-related heart rate increase achieved better performance. Patients with motor seizures showed significantly higher performance with ACC features, while HRV features were more informative for non-motor seizure detection.

**Significance:** This study validated the use of ACC and ECG-derived HRV for patient-independent seizure detection in long-term ambulatory data. ACC features were less generalizable overall, but improved performance for a subset of patients. Seizure type analysis revealed complementary strengths of autonomic and movement signals, highlighting the value of incorporating seizure type information into future multimodal systems.

## 1 Introduction

Epilepsy is a chronic neurological disorder characterized by recurrent seizures, which arise from excessive or hypersynchronous neuronal activity in the brain [1]. Depending on the affected regions, seizures can present with a wide range of symptoms, including changes in awareness, movement or behavior [2, 1]. Motor manifestations, such as jerking and stiffening, can occur in both focal and generalized seizures, and are often used clinically to recognize and classify seizures [3]. These characteristic movements form the basis of automated movement-based seizure detectors, many of which rely on accelerometry to capture abnormal motor activity [4, 5, 6].

Accelerometry is a widely used method for monitoring movement, with established applications in fall detection and activity tracking [7, 8]. Accelerometer sensors are embedded in consumer-grade wearable devices, such as fitness trackers, making them practical for chronic seizure monitoring in ambulatory environments [9, 10]. In epilepsy, accelerometry can characterize distinctive ictal movements: myoclonic and rhythmical clonic jerks produce low-frequency impulses in muscles (*<*6 Hz), while clonic muscle contractions are accompanied by high-frequency impulses (*>*10 Hz) [11]. Commercial accelerometry-based seizure detection systems often rely on these high-frequency rhythmic movements as a key principle to identify motor seizures [11], and their performance can be further improved when combined with other modalities such as electrodermal activity (EDA) and electromyography (EMG) [6, 12].

Although accelerometry is effective for detecting motor seizures, it is less suitable for seizures without motor manifestations. Most existing seizure detection devices are based on wrist- or arm-worn sensors and are designed primarily to detect convulsive seizures, particularly generalized tonic-clonic seizures, given their association with secondary physical injury and the risk of sudden unexpected death in epilepsy (SUDEP) [13, 14]. However, recognizing non-convulsive seizures is crucial for accurate diagnosis and effective treatment. These seizures often present with subtle non-motor symptoms, and patients may experience impaired awareness, leading to under-reporting [15]. Importantly, seizure-related autonomic changes, such as changes in heart rate and heart rate variability, can be used for seizure detection [16, 17]. This has motivated the inclusion of complementary signals to improve detection of a wide range of seizure types [18, 19, 20].

Despite the potential of multimodal seizure detection, several important gaps remain. Most prior studies have been conducted in hospital-based epilepsy monitoring units [18, 19, 21]. Many also include relatively small patient cohorts [22]. Although both accelerometry and autonomic signals have been shown to contribute to seizure detection, there is a lack of direct comparison between their relative value at the individual level. Our previous work demonstrated that electrocardiography (ECG)-derived heart rate variability features can support robust, patient-independent seizure detection in long-term ambulatory recordings [23]. The present study aimed to determine whether accelerometry provides complementary value to ECG-based seizure detection, in addition to validating the generalisability of seizure detection in a larger, unseen patient cohort. Using a large and diverse ambulatory video-EEG dataset, we evaluate multimodal seizure detection performance within a pseudo-prospective framework and analyze how individual patients benefit from motor versus autonomic modalities.

## 2 Methods

The dataset consisted of long-term ambulatory video-electroencephalography (vEEG) studies collected by Seer Medical (Australia) as part of standard clinical epilepsy assessment. A new cohort of patients was used compared to our previous work [23] (due to the availability of accelerometry), but the data was collected using the same wireless vEEG-ECG monitoring system with accelerometer sensor and clinical workflow. The wearable system included 21 scalp EEG electrodes and three ECG electrodes. An accelerometer sensor (ACC) was housed in the connector positioned on top of the head [24] and measured acceleration in three orthogonal axes. ECG and ACC data were sampled at 250 Hz. Patients wore the system continuously in their homes for 2-8 days, with EEG, ECG, ACC and video recorded throughout. Seizures were annotated by neurologists based on EEG and video, but only ECG and ACC were used as input to the detection algorithm. Figure 1 shows an example segment of ECG and ACC data over a seizure period.

**Figure 1:**
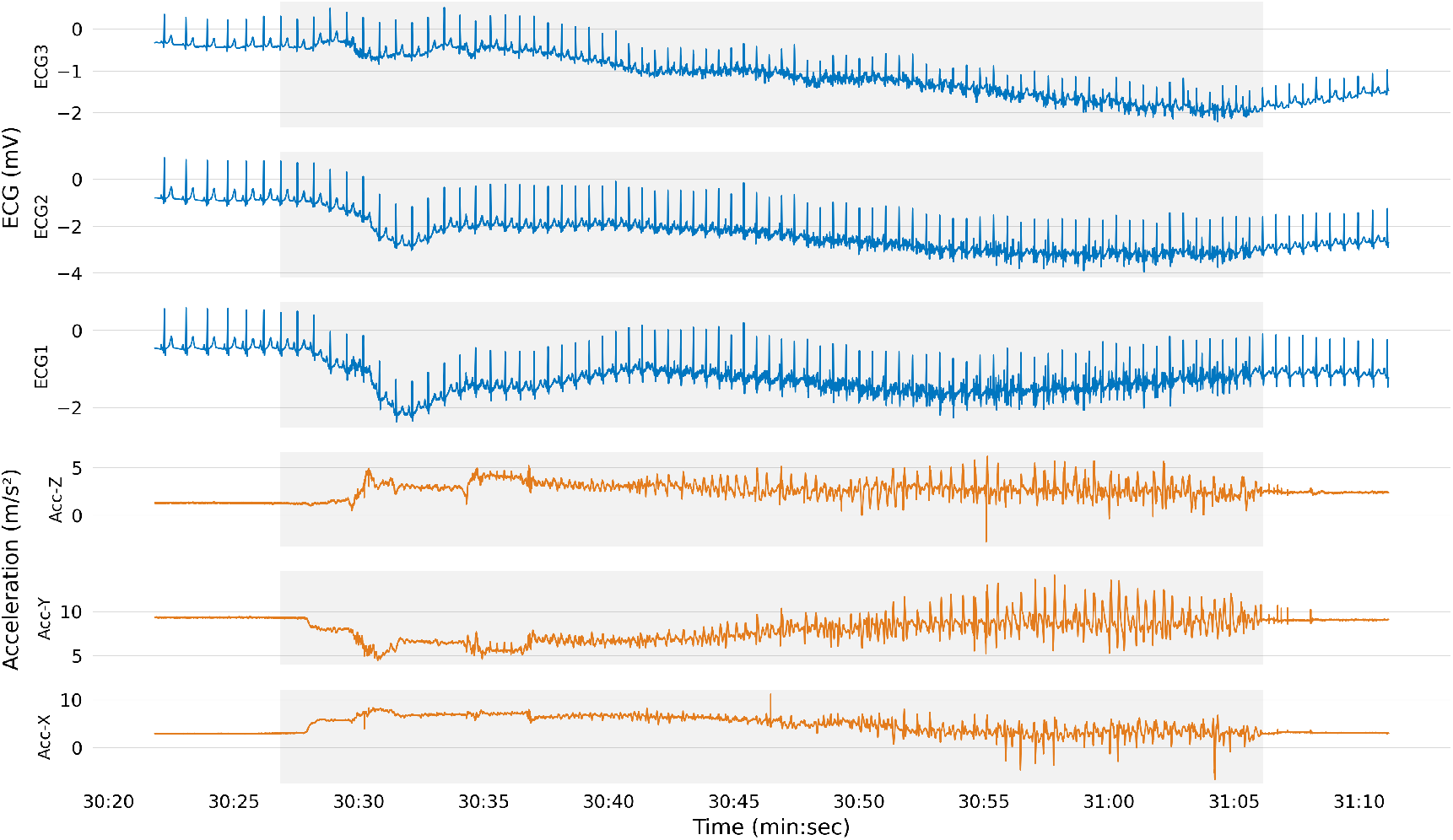
Raw ECG and accelerometry signals captured during a nocturnal seizure (duration highlighted in grey). Video showed the patient having non-purposeful limb movements. ECG: electrocardiography.

ECG preprocessing and feature extraction followed the pipeline established in our previous work [23]. Briefly, non-linear heart rate variability (HRV) features were extracted using Lorenz plot analysis in 50 s and 100 s windows at a 1 s step size [25]. Accelerometry data were preprocessed in non-overlapping 1 s windows. The extracted features included the mean and standard deviation of each axis and of the signal magnitude, the spectral energy of the magnitude signal via the fast Fourier transform and a previously validated signal quality metric [26]. ECG and ACC features were concatenated before classification.

A leave-one-patient-out approach was used to cross-validate the training set while establishing the machine learning approach. A logistic regression classifier was used to classify periictal (defined as 2 min before and after seizures) and interictal periods. After all development and training was completed, the classifier was frozen and performance was then assessed on a separate test set of patients not seen during training or validation.

Event-based performance metrics were used to evaluate the detector, to mimic real world clinical utility. Seizure detections were defined at the event level, where overlapping detections with a periictal segment were counted as true positives (TP), non-overlapping detections as false positives (FP), missed periictal events as false negatives (FN), and correctly identified interictal windows as true negatives (TN). Sensitivity (TP/(TP+FN)) and specificity (TN/(TN+FP)) were calculated along with two additional metrics: “Improvement over Chance” (IoC) and “Improvement over Chance of the Area Under the Receiver Operating Curve” (ΔAUC). IoC quantifies the gain in sensitivity over chance at a given detection rate. ΔAUC measures the improvement of the observed AUC over chance-level AUC, independent of threshold selection. These metrics addressed the fact that chance performance in event detection deviates from the conventional 0.5 baseline due to events overlapping consecutive detection windows, and so provided a less biased evaluation of event-level performance.

To assess whether the validation and holdout test sets were comparable in terms of patient and study characteristics, we performed the Mann-Whitney U test for continuous variables (patient age and study characteristics), and chi-square tests for sex, epilepsy type and ictal heart rate response, defined as an average *≥*50 bpm increase in maximum heart rate during seizures (also referred to as a “heart rate responder” for ECG-based seizure detection [25]). To evaluate factors affecting patient-level performance, we used the Mann-Whitney U test to compare patient groups in terms of sex, epilepsy type and whether they were a “heart rate responder”. The Pearson correlation test was used to compare whether individual performance was linked to patient age or the number of events.

Individual seizure types were not available at the level of each recorded event. However, every patient had an accompanying clinical report containing descriptions about reported and discovered events, and the neurologist’s summary of the ambulatory study. These reports were therefore used to extract likely seizure types at the patient level.

Seizure type terminology was identified using a predefined keyword dictionary covering focal motor, focal impaired awareness (IA), focal to bilateral tonic–clonic, generalized tonic–clonic (GTCS), absence, and myoclonic seizures. Keyword matching was case-insensitive and performed using regular expressions to allow for variations in phrasing and terminology.

For comparisons of seizure detection performance, seizure types were grouped into categories. Motor seizures included GTCS, focal motor, focal to bilateral tonic-clonic, and myoclonic seizures. Non-motor seizures included focal impaired awareness and absence seizures. Generalized seizure subtypes were grouped into GTCS and non-GTCS (absence or myoclonic). Focal seizure subtypes were grouped into focal motor (focal motor or focal to bilateral tonic-clonic), and focal impaired awareness only (focal impaired awareness without other motor seizures). Patients with no identifiable seizure types (*unclassified*) were excluded from group-level comparisons.

Differences in detection performance between seizure type groups were assessed using the Mann-Whitney U test. Individual performance was summarized using the Δ*AUC* metric, and comparisons were conducted for models trained using HRV-only, ACC-only and combined HRV+ACC feature sets.

## 3 Results

Patient demographics and dataset characteristics are summarised in Table 1. No significant differences were observed between the training and test cohorts in terms of patient or study characteristics. Each patient contributed between 1 and 35 periictal events (median: 3), with an average recording duration of 5.22 days and 93% of data available for analysis. Forty-six (59%) patients were diagnosed with focal epilepsy and the other 32 (41%) with generalized epilepsy. Eighteen patients were identified as heart rate responders (n=11 in training, n=7 in test).

**Table 1:** Summary of patient and study characteristics. Note that the number of periictal events might be less than the number of distinct seizures, as seizures clustered in time (within 5 minutes) were combined to a single event for detection. Values are reported for the whole cohort (n=78), validation set (n=47), and hold-out test set (n=31). *p*-values correspond to comparisons between validation and test sets, calculated using the Mann-Whitney U test for age and study characteristics, and chi-square tests for sex, epilepsy type and whether the patient is a heart rate responder.

|  | Total (n=78) | Validation (n=47) | Hold-out Test (n=31) | *p*-value |
| --- | --- | --- | --- | --- |
| **Patient Characteristics** |  |  |  |  |
| Sex |  |  |  | 0.49 |
| Male | 36 (46%) | 23 (49%) | 13 (42%) |  |
| Female | 41 (53%) | 23 (49%) | 18 (58%) |  |
| Other | 1 (1%) | 1 (1%) | 0 |  |
| Age (years) |  |  |  | 0.68 |
| median [IQR] | 21 [14-13.5] | 21 [12.5, 32] | 21 [12-32.75] |  |
| Epilepsy type |  |  |  | 0.31 |
| Focal | 46 (59%) | 25 (53%) | 21 (68%) |  |
| Generalised | 32 (41%) | 22 (47%) | 10 (32%) |  |
| Mean ictal heart rate changes |  |  |  | 0.76 |
| Responder | 18 (23%) | 11 (23%) | 7 (23%) |  |
| Non-responder | 60 (77%) | 36 (77%) | 24 (77%) |  |
| **Study Characteristics** |  |  |  |  |
| Study duration (days) |  |  |  | 0.26 |
| mean | 5.22 | 4.95 | 5.59 |  |
| Data availability (%) |  |  |  | 0.12 |
| mean | 93 | 92 | 96 |  |
| Periictal events per patient |  |  |  | 0.08 |
| median | 3 | 4 | 2 |  |
| Average seizure duration (min) |  |  |  | 0.97 |
| median | 1.18 | 1.16 | 1.27 |  |

There were no significant differences between the validation and hold-out test sets for any of the patient demographic variables. Specifically, age did not differ significantly between sets according to the Mann-Whitney U test, and chi-square tests showed no significant differences in the distribution of epilepsy type, sex or heart rate responder status. No significant differences were observed between the validation and hold-out test sets in study duration, data availability, periictal events per patient, or average seizure duration, as assessed using the Mann–Whitney U test.

Three feature sets were used to train and evaluate seizure detection models: HRV features only, ACC features only, and HRV and ACC features combined. The overall results are summarised in Table 2. Figure 2 shows the event detection performance for individual patients using the HRV and ACC combined model. Across all models, the majority of patients showed performance above chance, i.e., IoC*>*0. The combined HRV and ACC model yielded the highest improvement over chance AUC (ΔAUC=0.12) in the validation set, with a mean IoC of 0.24. However, in the hold-out tests, the model using only HRV features outperformed only ACC features and HRV and ACC features, suggesting that ACC features did not provide added value in the hold-out tests. ACC-only models showed the lowest performance overall, but still showed above-chance detection in over 74% of patients.

**Table 2:**
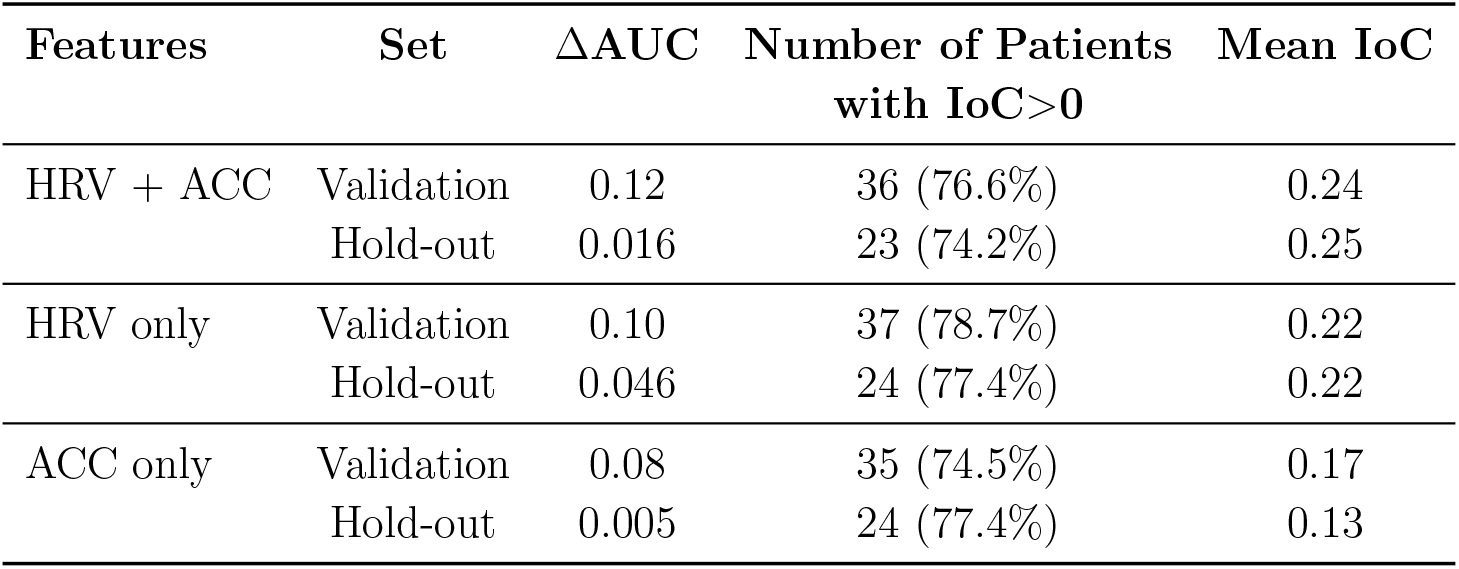
Seizure detection performance across feature sets and datasets. Performance is reported as the improvement over chance AUC (ΔAUC), and the improvement over chance (IoC). ΔAUC is a threshold-independent metric calculated by aggregating all events across patients, while IoC is threshold-specific and evaluated per patient. The proportion of patients with IoC*>*0 and the mean IoC value averaged across patients are reported. Results are shown for the validation and hold-out test sets across HRV+ACC, HRV-only, and ACC-only feature sets.

**Figure 2:**
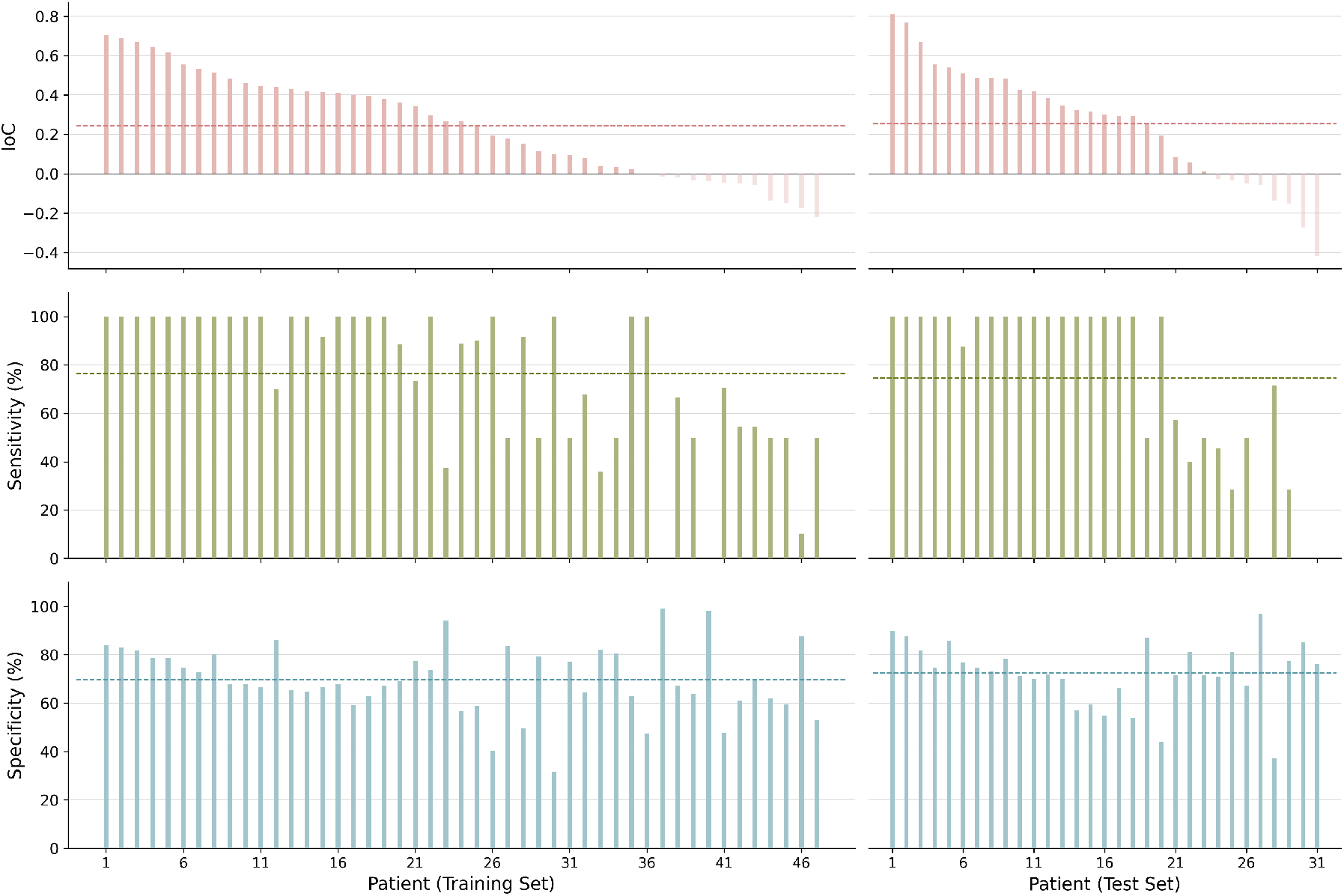
Event detection performance for leave-one-patient-out cross-validation (Training Set: n=47) and hold-out (Test Set: n=31) tests. Shown from top to bottom are: Improvement over chance (IoC) (red), sensitivity (green) and specificity (blue). Dashed lines show represent the population mean of each metric.

While the combined HRV and ACC model achieved the best performance on the validation data, its ability to generalize to the held-out data was limited, as evidenced by the drop in ΔAUC. In contrast, the model using HRV features showed more stable performance across validation and hold-out sets, with a relatively smaller drop in ΔAUC (0.054 compared to 0.104 change in performance). Performance using only ACC features was poorer, further suggesting the limited capability of ACC features for patient-independent seizure detection. Notably, the reduction in performance between validation and hold-out sets was more substantial in ΔAUC than in patient-averaged IoC. This reflects the fact that ΔAUC is influenced by the number of seizure and non-seizure events across patients, whereas IoC gives equal weight to each patient regardless of how much data they provide. The implications of these differences are discussed further in the Discussion section.

Figure 3 shows group-level performance using different feature sets (only HRV features, only ACC features and combined HRV and ACC features). The median patient performance was higher when all features were used. Patients were grouped based on sex, epilepsy type and whether they fit the heart rate responder criteria. Across all feature sets, patients with focal epilepsy showed better performance than patients with generalized epilepsy, though the difference was only statistically significant (p*<*0.05) for models using only HRV features and HRV+ACC features. In addition, heart rate responders, which are patients showing *>*50 bpm average heart rate difference during seizures, outperformed non-responders significantly in all feature sets.

**Figure 3:**
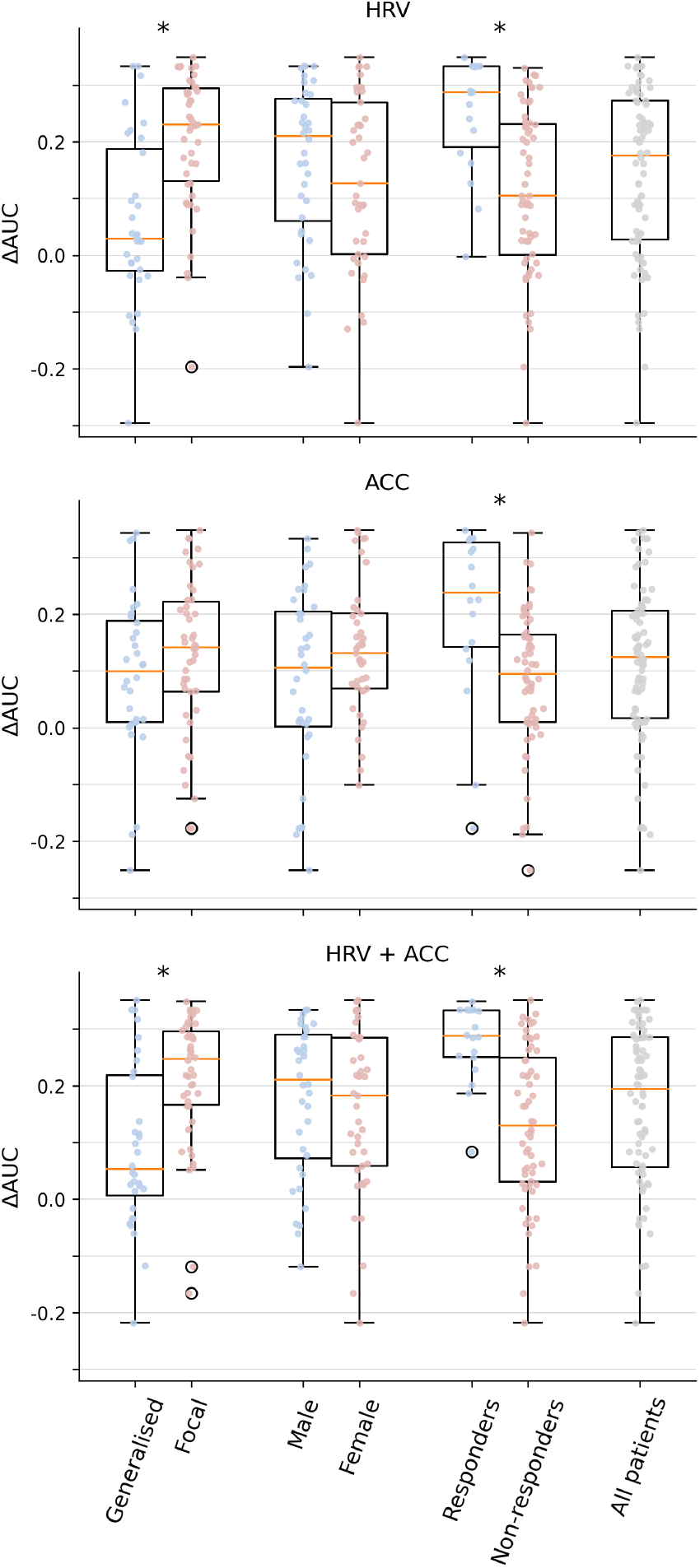
Improvement over chance AUC (Δ*AUC*) for patients, grouped by epilepsy type, sex, and average ictal heart rate (responders and non-responders). Each subplot shows results using (a) HRV features, (b) ACC features, (c) combined HRV and ACC features. Boxplot shows group-level distributions, with individual patients represented as coloured dots. Asterisks (*) denote statistically significant differences between the performance of subgroups (p*<*0.05).

Figure 4 shows the performance comparison using HRV features and ACC features by visualizing the difference in ΔAUC across patients using each feature set. Each line connects the ΔAUC value using HRV features (left) to that using ACC features (right). Lines with positive slope (blue) suggest that ACC features were more informative than HRV features at characterizing seizures, while lines with negative slope (red) suggest the opposite. Color intensity reflects the absolute value of the differences. Overall, across training and test sets, 33 out of 78 (42%) patients benefitted more from ACC features while 45 (58%) patients benefitted more from HRV features.

**Figure 4:**
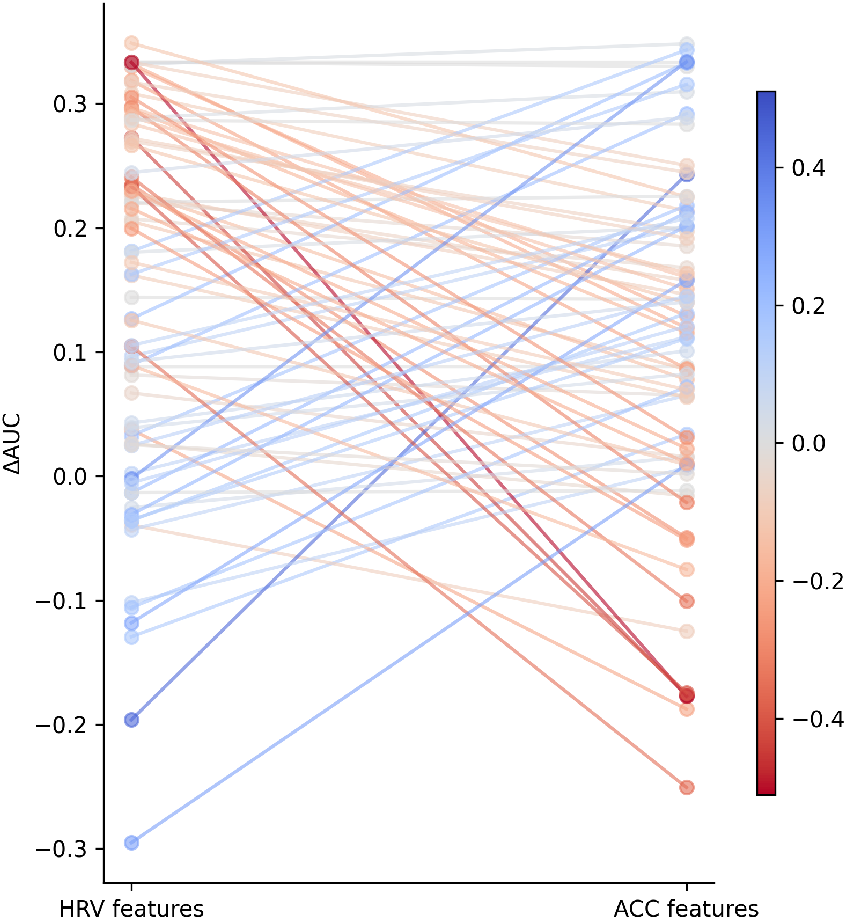
Improvement over chance AUC (Δ*AUC* for individual patients using HRV features vs. ACC features. Each line connects a patient’s Δ*AUC* scores using the two feature sets. Line colors indicate the direction and magnitude of the performance difference. Positive slopes (blue) suggest better performance using ACC features than using HRV features, while negative slopes (orange) suggest better performance using HRV features. Color intensity reflects the absolute value of the slope, i.e., the amount of performance difference.

Figure 5 summarises the number of patients with each seizure type, stratified by epilepsy type (focal or generalized). Of the 78 patients included across the validation and hold-out test sets, 20 (26%) had no identifiable seizure type descriptors in their reports, and were therefore labelled as unclassified, leaving 58 patients (74%) with seizure type classifications. Among these, 35 patients (60%) had focal epilepsy and 23 had generalized epilepsy (40%). Within the focal epilepsy group, 33 patients had focal impaired awareness seizures, and 9 had focal to bilateral tonicclonic seizures. No patients were identified as having focal motor seizures. Within the generalized epilepsy group, 5 patients had GTCS, 14 had absence seizures, and 8 had myoclonic seizures.

**Figure 5:**
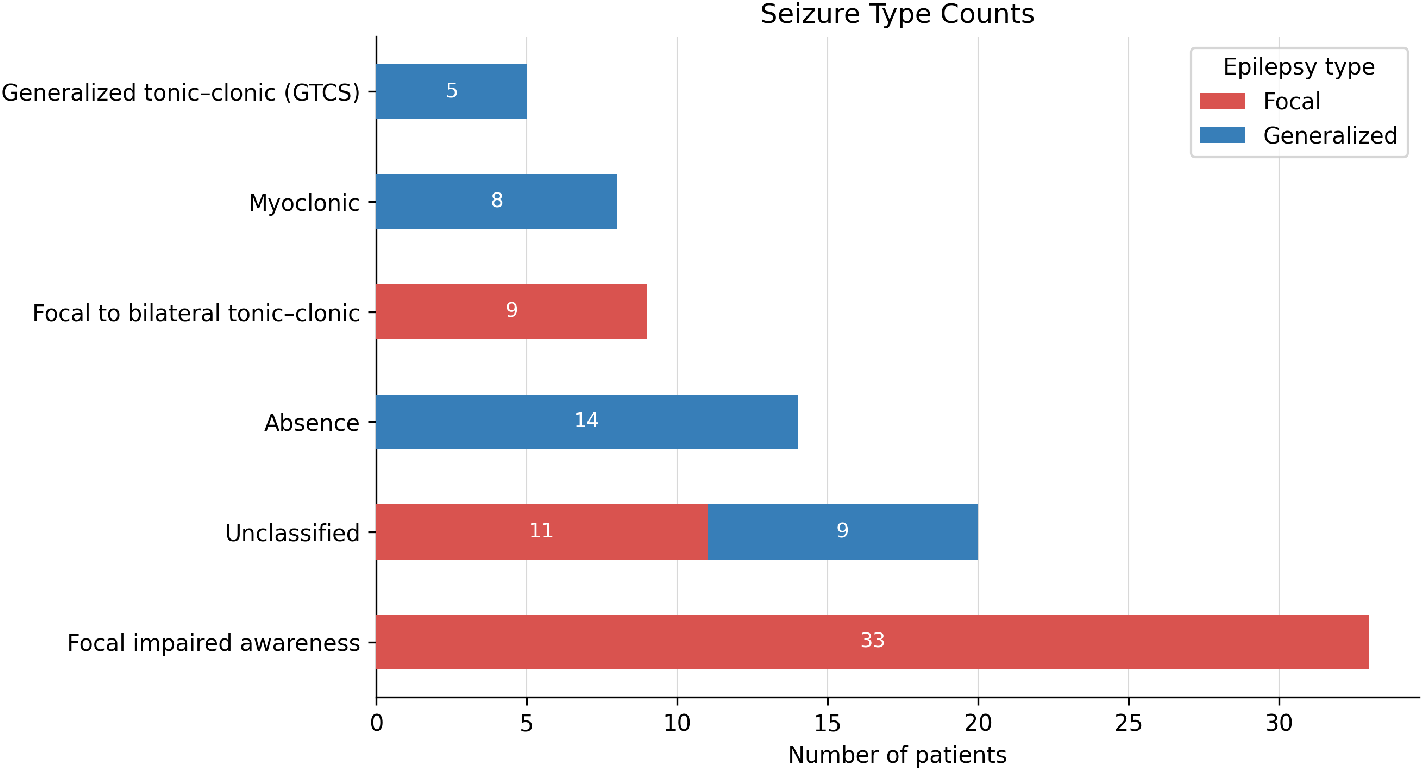
Horizontal stacked bars show the number of patients exhibiting each seizure type based on extraction from clinical reports. Colors indicate the epilepsy diagnosis, focal (red) or generalized (blue).

Figure 6 compares patient-level detection performance across seizure type groups using HRV-only, ACC-only, and combined HRV+ACC features. Detection performance as measured by ΔAUC across seizure type groups is summarised in Table B1 in Supplementary Material.

**Figure 6:**
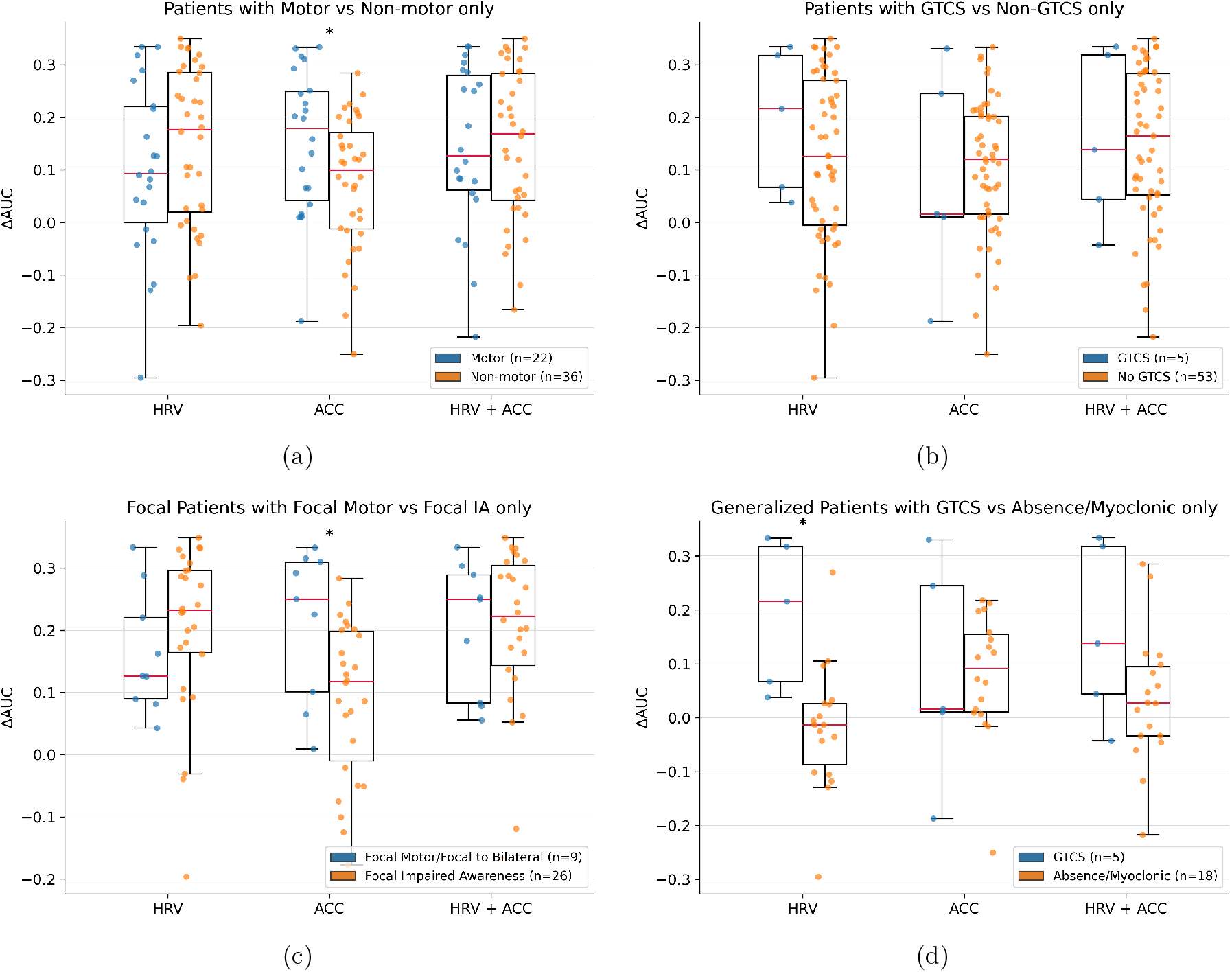
Comparison of patient-level seizure detection performance across seizure type groups. Boxplots show individual patient performance measured using the ΔAUC metric, for models trained with HRV-only, ACC-only, and combined HRV+ACC features. Scatter points represent individual patients, colored by groups. (a) Patients with motor seizures vs. patients with non-motor seizures only. Motor seizures include generalized tonic-clonic seizures (GTCS), focal motor, focal to bilateral tonic-clonic, and myoclonic; non-motor seizures include focal impaired awareness and absence seizures. (b) Patients with GTCS vs. patients with non-GTCS only. (c) Focal epilepsy patients with focal motor seizures vs. patients with focal impaired awareness (IA) seizures only. (d) Generalized epilepsy patients with GTCS vs. patients with absence/myoclonic seizures only. Asterisks (*) indicate statistically significant differences between groups based on Mann-Whitney U tests. Sample sizes for each group, *n*, are shown in the legend of each panel.

Patients with motor seizures outperformed those without motor seizures when using ACC-only features (*p*=0.04). In contrast, patients without motor seizures showed higher performance when using HRV-only features, although this difference was not statistically significant (*p*=0.33). With combined HRV+ACC features, performance improved relative to the weaker single modality in each group: motor-seizure patients showed gains relative to HRV-only models, whereas non-motor patients showed gains relative to ACC-only models.

When comparing patients with GTCS and those without (across both generalized and focal epilepsy), ACC-only performance was lower in the GTCS group (median ΔAUC=0.016 vs. 0.12). However, due to the very small number of GTCS patients (n=5), this difference was not statistically significant (*p*=0.89) HRV-only performance showed that GTCS patients showing a slightly higher median ΔAUC (0.22 vs. 0.13), but again this was not significant (*p*=0.32). Performance in the combined HRV+ACC model was similar between groups (ΔAUC=0.14 vs. 0.16, *p*=0.79).

Among patients with focal epilepsy, those with focal motor seizures (including focal to bilateral tonic-clonic seizures) performed significantly better than those with focal impaired awareness seizures when ACC-only features were used (*p*=0.017). The opposite trend appeared in HRV-only model, where patients with impaired awareness only showed better performance than those with focal motor seizures, although this was not statistically significant (*p*=0.21).

Among patients with generalized epilepsy, GTCS patients exhibited higher HRV-based performance than those with absence or myoclonic seizures only, with a large median difference (0.22 vs. −0.013, *p*=0.0025). However, ACC-only performance showed no clear advantage for GTCS, with absence or myoclonic only patients showing a slightly higher median ΔAUC, although this was not significant (*p*=0.86). With the combined HRV+ACC model, patients with GTCS showed higher median, but the difference was not significant (*p*=0.13).

## 4 Discussion

This study investigated seizure detection using ECG-derived heart rate variability and head-mounted accelerometry in a long-term ambulatory dataset of 78 patients with epilepsy. Unimodal (HRV-only and ACC-only) and multi-modal (HRV+ACC) seizure detection models were compared under both leave-one-patient-out cross-validation and hold-out test conditions. Seizure detection using HRV features alone yielded the most stable performance across validation and test sets, outperforming both the ACC-only model and the combined HRV+ACC model in the held-out data in terms of ΔAUC. While the combined model showed greater improvement over chance in cross-validation, its generalizability to unseen patients was limited. Patient-level analysis revealed marked heterogeneity: although 58% of the patients benefitted more from HRV features, 42% showed better detection performance with ACC features, highlighting the complementary nature of autonomic and movement signals.

Wearable seizure detection is increasingly recognized as a way to extend monitoring beyond clinical settings, with non-EEG signals offering a practical and less intrusive means of chronic, continuous recordings. Our findings provide further evidence by validating the utility of ambulatory physiological signals for seizure detection. Consistent with prior literature [18, 19], we observed that multimodal approaches can improve performance over unimodal methods, since the HRV+ACC model outperformed unimodal models during cross-validation. However, in pseudo-prospective hold-out testing, HRV features alone yielded more stable and generalizable performance, while the benefit of adding ACC did not translate across unseen patients. One possible explanation is that seizure-related autonomic changes captured by HRV are more consistent across individuals, while ACC features are more variable, depending on seizure type, motor manifestation, and background activity. Our patient-level analysis supports this interpretation: although some patients clearly benefited from ACC, others derived little added value beyond HRV. These findings suggest that, while multimodal integration holds promise, its utility may depend on individual patient profiles, underscoring the need for further personalization in seizure detection systems.

Hold-out performance appeared much lower than validation in terms of ΔAUC, but not in the percentage of patients with positive IoC or mean IoC. This discrepancy arises from differences in how these metrics are calculated. ΔAUC is computed by aggregating all events across patients, making it more sensitive to the number of events. Patients who contribute more data–whether through a larger number of seizures or longer recording durations resulting in more non-seizure windows–have a stronger influence on the overall ΔAUC. Although study characteristics such as study length, number of periictal events, and average seizure duration did not differ significantly between the validation and hold-out sets, small imbalances can amplify the influence of certain patients on the aggregated metric. Consequently, in a smaller hold-out test set, patients with lower performance or whose data is more difficult to classify can decrease the aggregated ΔAUC, even if most patients perform comparably well at the individual level. On the other hand, IoC is computed separately for each patient, and the mean IoC reported is averaged across patients, giving each individual equal weight regardless of the number of seizures or non-seizure segments they contributed. This makes IoC less affected by differences in data sizes across patients and better suited for assessing generalization at the patient level. However, patient-level IoC can be affected by small-sample bias [27]. For patients with few seizures, performance estimates are statistically less reliable, since one correctly or incorrectly detected seizure can shift the patient’s AUC and hence IoC considerably. Although averaging across patients mitigates the influence of any single unstable estimate, the mean and proportion of patients with positive IoC should still be interpreted with this limitation in mind.

Group-level comparisons revealed important sources of variability in detection performance. Consistent with previous work, patients with focal epilepsy achieved higher detection accuracy than those with generalized epilepsy, suggesting that autonomic alterations are more pronounced in focal seizures [23]. Patients with generalized epilepsy showed poorer performance, likely reflecting the fact that motor manifestations may not always translate into distinctive accelerometry patterns when using a head-mounted sensor. Furthermore, patients who exhibited pronounced ictal heart rate increases (i.e., heart rate responders) consistently outperformed non-responders across all feature sets. Because heart rate responder status was derived from heart rate changes across all seizures recorded from a patient, it can only be determined retrospectively rather than prospectively for unseen patients.

### 4.1 Comparison to Literature

Our previous study developed a seizure detection pipeline for ambulatory ECG data and introduced a pseudoprospective evaluation framework combining leave-one-patient-out cross-validation with adjusted event-level metrics [23]. Here, we applied the methodology to a larger and independent patient cohort, further validating and extending earlier findings. Using only HRV features, 78.7% of patients in the validation set and 77.4% of patients in the hold-out test set achieved above-chance performance, closely matching the previous results (74% and 78%, respectively). Mean improvement over chance (IoC) was also comparable between studies (0.22 vs. 0.20 in the hold-out test set). Furthermore, the present study expanded the hold-out cohort from 18 to 31 patients, further supporting the robustness and generalizability of ECG-derived HRV features for ambulatory seizure detection.

Movement-based seizure detection has long been a focus of wearable devices, particularly for generalized tonic-clonic seizures (GTCS), which are associated with characteristic rhythmic movements and high clinical risk. Previous epilepsy monitoring unit (EMU)-based studies report sensitivities around 90% with false alarm rates as low as 0.2/day for GTCS detection [28, 29, 5, 4]. However, these approaches were primarily developed to target GTCS and, therefore, performed poorly when applied to other seizure types. One study reported only 56% sensitivity for nocturnal motor seizures [21], while another detected 89.7% of GTCS but did not trigger alarms for 149 seizures of other types [5]. More recent EMU studies have been extended to broader seizure populations using multiple modalities. Their results using only ACC yield AUC-ROC values of 0.783 [18] and 0.673 [19]. In comparison, the current ACC-only model achieved AUCs of 0.75 (validation) and 0.67 (hold-out), closely matching the reported performance of in-patient settings [18, 19]. Differences in evaluation frameworks, particularly our use of ΔAUC to account for chance performance under event-based windowing, as well as differences in sensor placement (top of head in our study instead of wrist in previous studies), limit direct comparison.

To address the challenge of detecting non-GTCS seizures, multimodal approaches have been explored to combine autonomic and motor signals. Munch Nielsen et al. [22] reported sensitivities of 67-78% and false alarms of 40-48/day when combining ECG and sternum-mounted ACC in three patients. Similarly, van Andel et al. [21] analyzed 86 nocturnal motor seizures from 23 patients and achieved 71% sensitivity and 5.9 false alarms per night (8 hours) using combined heart rate and ACC. Larger EMU-based studies by Yu et al. and Tang et al. [18, 19] found that combining ACC with blood volume pulse (BVP, a peripheral measure of cardiac activity via photoplethysmography [30]) improved leave-one-patient-out cross-validation performance to AUC values of 0.798 and 0.721, respectively. Yu et al. [18] reported sensitivity of 83.9% and FPR of 35.3%. Our combined HRV+ACC model achieved an AUC of 0.78 in validation. Threshold-specific metrics yielded TPR of 72.7% and FPR of 31.1%, representing lower sensitivity but slightly improved specificity relative to Yu et al. [18]. These results were obtained in a long-term ambulatory dataset rather than in the controlled EMU environment, highlighting both the potential and the challenge of translating multimodal seizure detection into real-world settings. The device used was not as compact or socially invisible as wristwatch-style wearables or adhesive patches, which may have influenced both data quality and patient behavior. These could contribute to variability in performance, and future work should investigate how device form factor and wearability influence performance.

### 4.2 Clinical Considerations

Most seizure detection studies to date have focused on real-time or low-latency detection, especially of GTCS, with the aim of alerting clinicians or caregivers to intervene [5, 31, 4, 6, 12]. Although some approaches report encouraging results, many are still hampered by false alarm rates that limit their clinical utility in this context [21, 18]. In contrast, other applications of wearable seizure detectors aim to provide a more objective method of documenting seizures to complement or improve self-reported seizure diaries [32, 33], which are often unreliable [15, 34, 35, 36]. Improving the accuracy of seizure documentation could directly enhance clinical decision making and ongoing treatment.

The present work adds clinical relevance by analyzing a relatively large cohort of patients who were continuously monitored in the home environment, performing daily activities, rather than in a highly controlled EMU environment. Seizures were verified by neurologists, and patients were not extensively screened for seizure or epilepsy type. This means that the evaluation more closely resembles a realistic scenario in which a patient-independent seizure detector is applied prospectively to unseen patients without prior training data. Key elements of the pseudo-prospective framework, such as threshold selection and hold-out testing, were designed to approximate this real-world application.

Building on previous work using ECG-derived features alone, the current study added ACC features, given their proven value in identifying seizure-related motor symptoms [11]. Similar to the challenge of using ECG to distinguish seizure-related autonomic changes from non-seizure changes, the challenge of using ACC is in differentiating seizure-related movement patterns from normal everyday activities. This is arguably more difficult, since ACC signals are highly variable and even normal activities or behaviors are difficult to characterize [37]. However, the presented findings showed that ACC provided complementary information and contributed to seizure detection in ambulatory settings for many patients. The contributions of the ACC signal were similar to previous studies, despite using an unconventional sensor location on the head rather than the wrist or limbs, which is more common in the literature [11].

### 4.3 Seizure Type Analysis

Seizure type information was systematically extracted from clinical reports and examined how seizure semiology, classified at the patient level, relates to detection performance using autonomic (HRV) or movement (ACC) signals. This analysis linked patient performance to seizure semiologies, and examined how contributions of the signal modalities vary across seizure categories.

When comparing patients with GTCS to those without GTCS (across both focal and generalized epilepsy), performance differed across modalities. GTCS patients showed substantially higher HRV-based performance. However, ACC-only performance was higher in the non-GTCS group. In this analysis, the non-GTCS group is not limited to non-motor seizure types, but includes patients with other motor semiologies, such as focal motor and focal to bilateral tonic-clonic seizures, which are associated with autonomic and movement changes [38, 39]. The inclusion of these motor seizure types likely contributes to the higher performance observed in the non-GTCS group.

Within generalized epilepsy, patients with GTCS outperformed those with absence or myoclonic seizures only when using HRV or combined HRV+ACC features, while ACC-only performance showed a slight advantage for absence or myoclonic seizures. This suggests that GTCS was relatively easier to detect when compared against other generalized seizure types, particularly when autonomic features were used.

The small size of the GTCS group (n=5) limits statistical power and make the results particularly sensitive to inter-patient variability. This may partly account for the inconsistent patterns observed across modalities. GTCS are relatively uncommon in ambulatory monitoring, where antiseizure medications are not withdrawn for safety reasons. In contrast, inpatient monitoring usually involves medication tapering to provoke convulsive events under clinical supervision [40]. GTCS recorded in ambulatory environments may therefore differ from those observed in controlled clinical settings. Furthermore, the ACC features used in this study were designed to capture general activity magnitude and fluctuations, rather than the high-frequency patterns that characterize convulsive movements, which may have limited ACC-related performance gains in the GTCS group.

While GTCS did not appear to benefit from ACC features, the advantage of ACC was more apparent when comparing patient groups defined by motor involvement. In both the motor vs. non-motor and focal motor vs. impaired awareness comparisons, ACC-only performance was significantly higher in the motor groups. Conversely, HRV-based detection tended to perform better in non-motor seizures, although the differences were not significant. The combined HRV+ACC model improved median performance relative to the weaker single modality in each group, suggesting that the two modalities contribute to complementary information, whose relative importance depends on seizure semiology.

These findings demonstrate that wearable-based seizure detection performance is influenced not only by signal modality, but also by seizure semiology. Different seizure types evoke distinct combinations of autonomic and motor responses, which determine how effectively they are captured by HRV and ACC features. This highlights the importance of integrating seizure type information into future multimodal detection frameworks, so that algorithms can better adapt to the physiological characteristics of different seizure types. The complementary strengths of HRV and ACC highlight the potential of multimodal approaches for improving generalizability across different seizure types.

### 4.4 Limitations and Future Work

While our data set was relatively large compared to previous work, the number of seizures per patient was limited, reducing the statistical power for patient-level analyses. This limitation, however, mirrors real-world clinical variability and aligns with the goal of developing models that generalize to patients with infrequent seizures. Future work could benefit from using open-source datasets, which can provide access to larger and more diverse cohorts [41]. In addition, accelerometry was recorded using a head-mounted device, which may capture some seizure-related movements but is less sensitive to limb activity than wrist-worn sensors. This placement likely influenced the performance of the modality.

A key limitation of the seizure type analysis is that seizure types were inferred at the patient level based on the clinical reports, rather than annotated for each individual seizure. As a result, the comparisons reflect the performance of patient groups with particular seizure types, not the detection performance for specific seizure events. Differences in seizure frequency, number of recorded events, and intra-patient variability may, therefore, influence group-level trends. More precise event-level annotation of seizure semiology would enable a more direct assessment of each modality to different seizure types.

Future work should incorporate event-level semiology labels, so that performance can be evaluated per seizure type rather than only at the patient level. This would enable models to adapt to physiological patterns associated with different seizure semiologies, thus improving the robustness and generalizability of seizure detection systems.

Our results highlight a key challenge in current wearable seizure detection research, which is data scarcity and imbalance. Many prior studies included only a handful of patients with usable seizure data, and even in larger cohorts, the number of seizures is small considering the inter-patient variability present in people with epilepsy. The scarcity of seizure events, in addition to the significant imbalance with non-seizure data, can limit the complexity of models that can be effectively trained, as well as the statistical robustness of results obtained from more complex approaches. Unlike EEG, where ictal changes can be directly observed in the signal in electrographic seizures, autonomic or movement alterations are indirect manifestations of seizures. Periictal autonomic or movement alterations are not present in all people with epilepsy, and even within the same patient may accompany some seizures but not others. For example, autonomic changes may be prominent in some seizures but absent in others within the same patient, and are less commonly observed in shorter, less severe seizures [16, 42]. Similarly, seizures characterized by impaired awareness or unresponsiveness may produce minimal motor signs [43]. As a result, the signals available to the model do not always reflect the underlying ictal process, meaning that seizures can vary widely in terms of how strongly they exhibit relevant physiological changes.

The combination of data scarcity and the indirect nature of non-EEG signals may explain why multimodality does not automatically improve performance at the patient level. The small sample size limits the ability of the model to capture the full variability of seizure manifestations across patients. In addition, autonomic or movement changes are not consistently present in all seizures or patients, meaning that the additional modality may not always contribute seizure-related information. Hence, a simple combination of modalities may be insufficient, and more sophisticated fusion strategies such as patient-specific or adaptive weighting may be required [44, 45]. Future research may also benefit from advanced feature engineering and modelling approaches that are capable of capturing complex seizure-related patterns in accelerometry. Deep learning methods could potentially extract subtle and patient-specific motor signatures that complement autonomic changes. However, training such models relies on large and well-annotated datasets that capture the diversity of seizure types, symptoms and patient contexts. It is otherwise difficult to drive clinically meaningful progress in applying machine learning or deep learning approaches to uncover subtle biomarkers in non-EEG data. Without sufficient examples of seizures across patients, modalities, and seizure types, models may fail to generalize to new patients, producing inconsistent performance and limiting their reliability in real-world, ambulatory settings.

A key focus of our future work will be to reduce the false detection rate by improving the ability of models to distinguish seizure-related physiological changes from normal fluctuations. This could be approached by incorporating additional information, such as patient-specific circadian patterns or activity levels, to better separate noise from seizure-related signals [46]. Adding other sensor modalities, such as electrodermal activity, also holds promise, as these signals may provide complementary information that helps reduce false alarms. Finally, enhancing model interpretability will be crucial, as it could provide insight into how non-EEG signals support detection across different patient subgroups or seizure characteristics, ultimately guiding both model refinement and more personalized applications.

Epilepsy is a complex disorder that affects not only the brain but the entire body, and seizures represent a process in the brain that produces various downstream physiological effects. Multimodal seizure detection using multiple wearable sensors offers a way to capture these systemic and heterogeneous manifestations more thoroughly. This helps us better understand how seizures impact the body and ultimately brings us closer to clinically meaningful applications.

## Data Availability

All data produced in the present study are available upon reasonable request to the authors.

## Ethics Statement

This study was performed in accordance with the Declaration of Helsinki. This human study was approved by the St Vincent’s Hospital Human Research Ethics Committee (HREC 009.19). All parents, guardians or next of kin provided written informed consent for minors to participate in this study. All adult participants provided written informed consent to participate in this study.

## Appendix A Statistical Tests

**Table A1:**
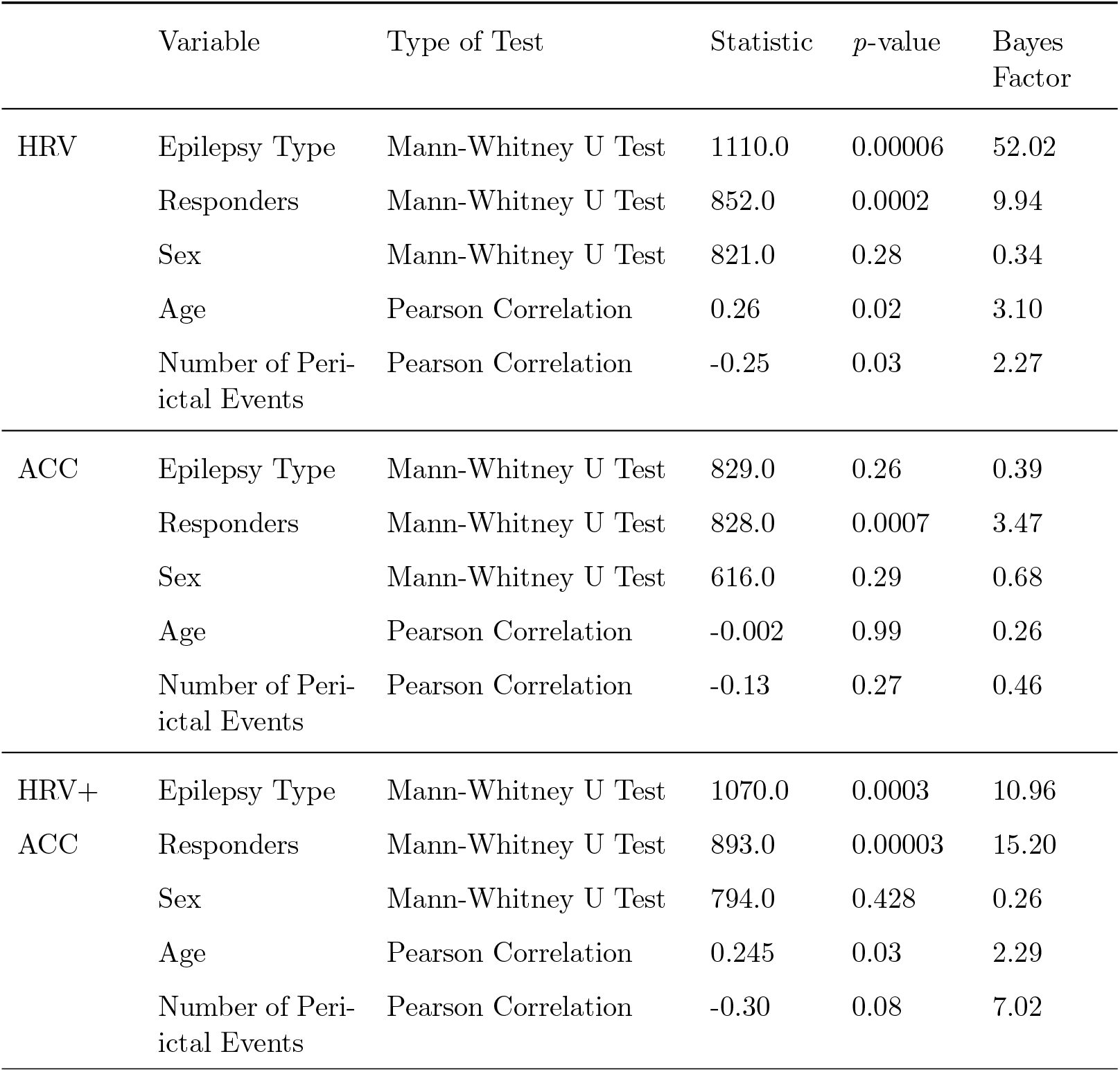
Statistical test results examining the effects of patient and study characteristics on model performance. Results are presented for three feature sets: HRV, ACC, and HRV+ACC.

## Appendix B Detection Performance by Seizure Type

**Table B1:** Detection performance (median ΔAUC) across seizure-type groups for HRV-only, ACC-only, and combined HRV+ACC models. *p*-values from Mann–Whitney U tests.

| Comparison | Group | N | HRV |  | ACC |  | HRV+ACC |  |
| --- | --- | --- | --- | --- | --- | --- | --- | --- |
| | | | Median ΔAUC | $p$ | Median ΔAUC | $p$ | Median ΔAUC | $p$ |
| Motor vs. Non-motor only | Motor | 22 | 0.09 | 0.33 | 0.18 | 0.04 | 0.13 | 0.98 |
|  | Non-motor | 36 | 0.18 |  | 0.099 |  | 0.17 |  |
| GTCS vs. Non-GTCS only | GTCS | 5 | 0.22 | 0.32 | 0.016 | 0.89 | 0.14 | 0.79 |
|  | Non-GTCS | 53 | 0.13 |  | 0.12 |  | 0.16 |  |
| Focal motor vs. IA only | Focal motor | 9 | 0.13 | 0.21 | 0.25 | 0.017 | 0.25 | 0.89 |
|  | IA-only | 26 | 0.23 |  | 0.22 |  | 0.22 |  |
| GTCS vs. Abs/Myoclonic only | GTCS | 5 | 0.22 | 0.0025 | 0.016 | 0.86 | 0.14 | 0.13 |
|  | Abs/Myoclonic | 18 | -0.013 |  | 0.092 |  | 0.027 |  |

## Notes

### Competing Interest Statement

The authors have declared no competing interest.

### Funding Statement

This study did not receive any funding.

